# Protection of vaccine boosters and prior infection against mild/asymptomatic and moderate COVID-19 infection in the UK SIREN healthcare worker cohort: October 2023 to March 2024

**DOI:** 10.1101/2024.09.11.24313477

**Authors:** Peter D Kirwan, Sarah Foulkes, Katie Munro, Dominic Sparkes, Jasleen Singh, Amanda Henry, Angela Dunne, Jean Timeyin, Sophie Russell, Jameel Khawam, Debbie Blick, Ashley D Otter, Nipunadi Hettiarachchi, Michelle D Cairns, Christopher H Jackson, Shaun Seaman, Colin S Brown, SIREN Study Group, Ana Atti, Jasmin Islam, Andre Charlett, Daniela De Angelis, Anne M Presanis, Victoria J Hall, Susan Hopkins

## Abstract

**Objective:** To estimate the protection of COVID-19 vaccine boosters against mild/asymptomatic and moderate SARS-CoV-2 infection over a 6-month period of XBB.1.5 and JN.1 variant circulation.

**Design:** Multi-state model applied to cohort study, adjusted for vaccination, prior infection, and demographic covariates.

**Setting:** National Health Services (NHS) hospitals in the UK.

**Participants:** Healthcare worker cohort including 2,867 eligible people with >6 months since a previous booster who tested fortnightly for SARS-CoV-2 between October 2023 and March 2024 and completed symptoms questionnaires.

**Main outcome measures:** Vaccine effectiveness (VE) of vaccine boosters received in October 2023 (baseline: booster >6 months prior), and durability of protection from a recent (past 6 months) previous infection (baseline: last infection >2 years prior) against mild/asymptomatic and moderate SARS-CoV-2 infection. Mild symptoms included acute respiratory symptoms for <5 days, moderate symptoms included influenza-like illness, acute respiratory symptoms for 5+ days, or sick-leave. VE and acquired protection were estimated from the multi-state model as: 1 – adjusted hazard ratio.

**Interventions:** Receipt of a COVID-19 bivalent original/BA.4-5 or monovalent XBB.1.5 booster during October 2023.

**Results:** Half of eligible participants (1,422) received a booster during October 2023 (280 bivalent, 1,142 monovalent) and 536 (19%) had at least one PCR-confirmed infection over the study period. For the monovalent booster, VE against infection was 44.2% (95% confidence interval 21.7 to 60.3%) at 0-2 months, and 24.1% (-0.7 to 42.9%) at 2-4 months post-vaccination, with no evidence of protection by 4-6 months. For the bivalent booster, VE against infection was 15.1% (-55.4 to 53.6%) at 0-2 months and 4.2% (-46.4 to 37.3%) at 2-4 months. VE (monovalent or bivalent) against moderate infection was 39.7% (19.9 to 54.6%), and against mild/asymptomatic infection was 14.0% (-12.1 to 34.0%). Controlling for vaccination, compared to those with an infection >2 years prior, infection within the past 6 months was associated with 58.6% (30.3 to 75.4%) increased protection against moderate infection, and 38.5% (5.8 to 59.8%) increased protection against mild/asymptomatic infection.

**Conclusions:** Monovalent XBB.1.5 boosters provided short-term protection against SARS-CoV-2 infection, particularly against moderate symptoms. Vaccine formulations which target the circulating variant may be suitable for inclusion in seasonal vaccination campaigns among healthcare workers.

**Funding:** UK Health Security Agency, Medical Research Council, NIHR HPRU Oxford, and others.

## Introduction

A year on from the COVID-19 public health emergency of international concern being declared at an end by the World Health Organisation [1], SARS-CoV-2 infection continues to result in clinically relevant illness and hospitalisation [2]. In the UK, most of the population has now attained hybrid immunity against SARS-CoV-2 infection from vaccination and prior infection [3]. Healthcare workers (HCWs), particularly those of ethnic minority backgrounds, experienced greater risk of infection during the pandemic [4,5]. Given the nature of their role, this group remains at potentially greater risk of infection and continues to be prioritised for annual autumn COVID-19 vaccine ‘boosters’, offered alongside seasonal influenza vaccines in the UK [6,7] (with co-administration of these vaccines having no significant impact on long-term immunogenicity [8]).

Studies among UK healthcare workers have found the additional protection against infection provided by previous COVID-19 mRNA boosters to be modest and short-lived compared to protection acquired from previous infection [9,10]. Nonetheless, when optimally timed, booster vaccines can help to reduce the burden of infection on the healthcare service and may lessen symptoms at the individual level [11–13].

In autumn 2023, during a period of XBB.1.5 variant circulation, mRNA boosters were offered to all UK healthcare workers; these were the third annual boosters offered to this group following previous boosters in autumn 2022 and 2023. The boosters administered were initially the bivalent original/BA.4-5 formulation, offered from September to mid-October 2023. These were superseded by the monovalent XBB.1.5 formulation during October 2023, providing an opportunity to investigate the relative effectiveness of updated vaccine formulations against infection and symptom severity.

The aims of this study were to estimate the overall and differences in protection of the bivalent and monovalent boosters administered during October 2023 against mild/asymptomatic and moderate infection over a 6-month period.

## Methods

### Study design and data sources

The SIREN study, run by the UK Health Security Agency (UKHSA), is a prospective cohort study of National Health Service (NHS) HCWs who have been followed continuously since June 2020. At enrolment participants completed a baseline questionnaire and provided nasal swabs for SARS-CoV-2 polymerase-chain reaction (PCR) testing, and serum samples for anti-SARS-CoV-2 antibody testing [14]. During the autumn/winter 2023/24 season (September 2023 to March 2024), enrolled participants provided fortnightly swabs and underwent regular antibody testing for SARS-CoV-2, and completed fortnightly symptom questionnaires.

### Participants

Inclusion criteria for this study were: >6 months having elapsed since receipt of a previous COVID-19 booster, and at least two SARS-CoV-2 PCR tests contributed during the follow-up period (1st October 2023 to 31st March 2024). Participants who had received more than five vaccine doses before October 2023 were excluded. Questionnaire responses, PCR and antibody test results (including from outside the study), and information on vaccination were collected centrally by UKHSA [14].

### Covariates

Demographic covariates included: age, gender, ethnicity, region of residence, occupational setting, staff type, medical conditions, and household status (Table 1). Linked testing and booster vaccination data included: PCR and antibody dates and results, vaccination date, and formulation (vaccine type).

Participant follow-up was categorised by time since previous infection (i.e. a time-varying covariate), using either the most recent date of a PCR positive test or anti-N positive result (Roche Elecsys anti-SARS-CoV-2 nucleocapsid (anti-N)). To be assigned the status of “infection naïve”, participants were required to have had no history of positive results (PCR or antibody) prior to the start of the analysis period, and to be confirmed as anti-N negative on a serum sample provided between September 2023 to March 2024. Participants for whom infection status at the start or during the analysis period could not be assigned were excluded from analyses (n = 703; Figure 1). A sensitivity analysis including these participants was conducted, with VE estimates very similar to the main analysis (Figure S6).

**Figure 1.**
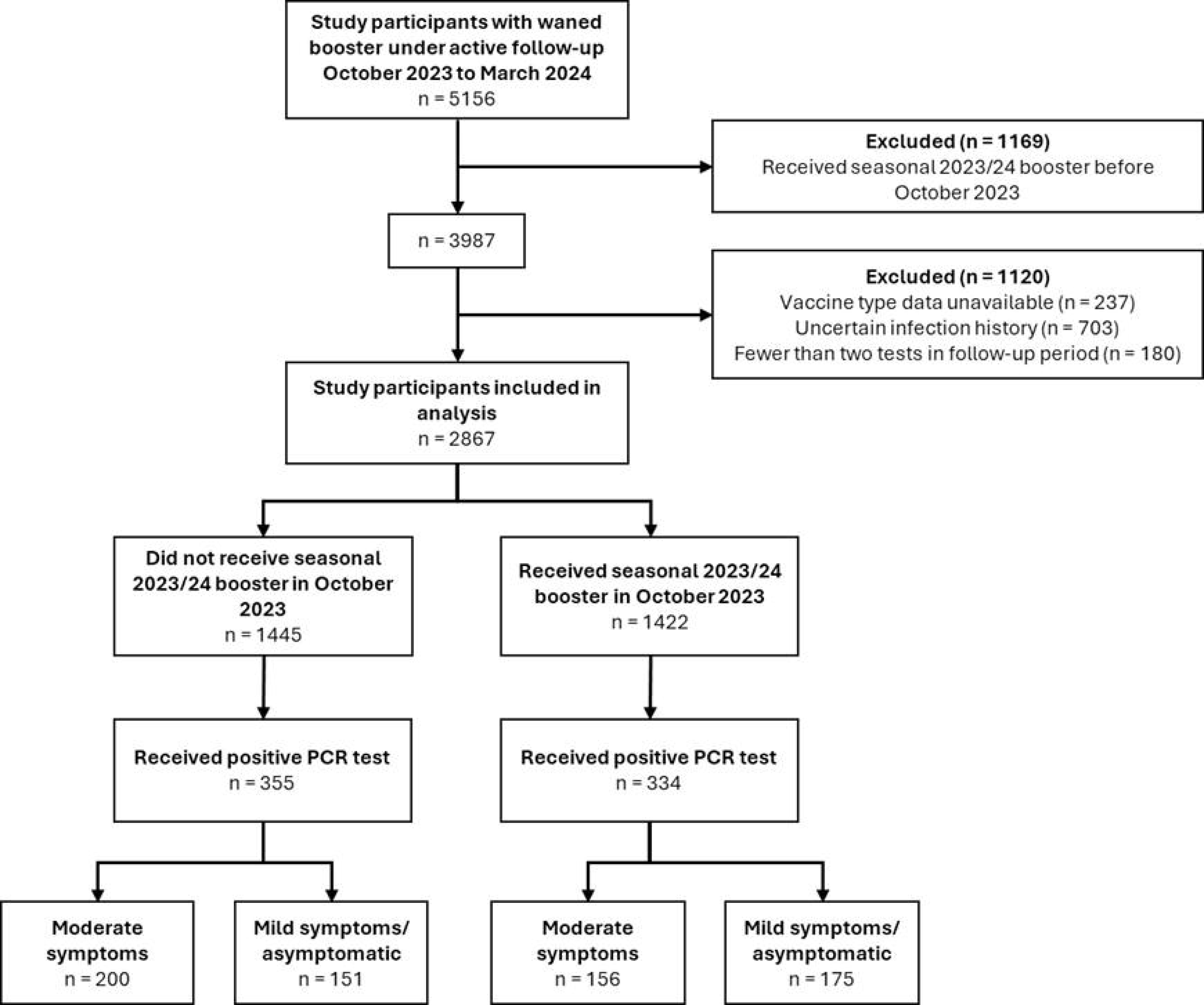

Questionnaires completed within a 14-day window of a positive PCR test were used to distinguish between illness with moderate (clinically relevant) symptoms and mild/asymptomatic COVID-19. Moderate symptoms were defined as influenza-like illness (ILI), or acute respiratory illness (ARI) lasting 5 or more days, or any sick leave. Episodes without symptoms, or with acute respiratory symptoms lasting <5 days, or with non-acute respiratory symptoms were defined as mild/asymptomatic illness. A full definition is included in the supplementary materials.

### Representativeness

All NHS hospitals and health boards in the UK were invited to join SIREN [14]. Study participants enrolled for autumn/winter 2023/24 broadly reflect the demography of UK HCWs, although with reduced representation of younger HCWs (aged <35) compared to previous studies of this cohort [15].

### Bias

The fortnightly testing regime minimised bias in detection of SARS-CoV-2 infection, and the statistical methodology further controlled for gaps in testing. To minimise recall bias, only symptoms reported within a 14-day window of a positive PCR test were considered.

### Censoring and late entry

Participants with a waned booster (>6 months since receipt of a previous booster) joined the study analysis at the date of their first PCR test after 1st October 2023. Participants with fewer than 24 weeks (6 months) since receiving a previous booster only entered the study once 24 weeks had elapsed. Participants were right-censored at their last recorded PCR test. As with previous analyses of this cohort, we allowed for deferred study entry and re-entry, with participants entering or re-entering the cohort 90 days after the date of a positive PCR test. Testing dates were pre-determined upon study enrolment with infection status only assumed to be known at the time of testing, and infection times assumed unknown. In principle, infections may have been undetected, especially if participants missed a testing appointment, so an appropriate statistical methodology was chosen to account for this.

To account for dependence between the month of vaccination and vaccine type, only boosters received during October 2023 (when both the bivalent and monovalent vaccine formulations were available) were considered. Participants who received their booster after October 2023 were right-censored at the date of vaccination and only contributed un-boosted follow-up time.

### Statistical analyses

Crude PCR positivity rates were calculated as the number of detected PCR positive results per 10,000 person-days of follow-up, with study re-entry. An exact Poisson method was used to calculate 95% confidence intervals [16].

Hazards associated with infection and mean time spent in the PCR positive state were estimated using multi-state models with covariate adjustment on month of study, vaccination status, time since previous infection, age group, gender, region, household status, ethnicity, staff type, occupation/setting, medical risk group, and COVID-19 patient contact. Eight multi-state models were used in total (M1 to M8), with each model assessing different aspects of VE, adjusted for all other covariates (M1: binary vaccination status; M2: time since vaccination; M3: vaccine type; M4: time since vaccination, by vaccine type; M5-M8: as for M1-M4, by symptom status). We compared the multi-state model estimates to those from a corresponding Cox proportional hazards model. The key difference between these two models is that multi-state models account for the incomplete information about infection times and potential for missed infections, but make stronger assumptions about the baseline infection rate, while the Cox proportional hazards models assume infection times are exact and complete, but do not make assumptions about the baseline rate.

We assessed model fit by comparing expected and observed numbers in each state over time, and undertook variable selection using Akaike information criterion values, and likelihood-ratio tests. Stratification was used to account for non-proportionality in the Cox models, and piecewise-constant baseline hazards by calendar month were used to account for dependence of the transition rates on time in the multi-state models. Vaccine effectiveness (VE) and acquired protection were estimated from the multi-state model as: 1 – adjusted hazard ratio [15]. We used 2+ years as the baseline for the time since previous infection due to the small size of the immunological baseline group (no prior SARS-CoV-2 infection). Full details of these models are included in the supplementary materials.

### Model implementation

Statistical models were implemented using R v.4.4.1 (R Foundation, Vienna, Austria) and the R package msm was used to fit the multi-state models [17].

### Patient and public involvement

The SIREN study has an active Participant Involvement Panel (PIP), co-delivered with the British Society for Immunology. The SIREN PIP meets regularly to consult participants on proposed study developments, recent outputs, and to obtain feedback on study delivery. The multimethod cohort retention programme is informed by the PIP and wider participant feedback is collected continuously through cross-sectional surveys.

## Results

### Population characteristics

A total of 2,867 participants were included in the analysis. The majority (81%) were female, aged between 35 and 64 (91%), and of White ethnicity (86%). A quarter (26%) reported a medical condition, including 2.4% with immunosuppression. Participants included HCWs from all disciplines, with the largest groups being nurses (33%), administrative/executive staff (16%), and doctors (13%) (Table 1). SIREN recruited HCWs from every UK region, but this analysis excluded Wales and Northern Ireland (n = 237 participants) where information on vaccine type was unavailable (Figure 1).

At study entry in October 2023, two-thirds (67%) of participants had a PCR-confirmed infection within the past 2 years, and a further 27% had their previous PCR-confirmed infection >2 years prior. The remaining 166 individuals (6%) had no PCR-confirmed evidence of infection and were confirmed as naïve based on their antibody results. During October 2023, 50% of participants (1,422) received a booster vaccination, of whom 280 (20%) received the bivalent original/BA.4-5, and 1,142 (80%) received the monovalent XBB.1.5 formulation (Table 1).

### Crude PCR positivity rates

Over the 6-month follow-up period, 536 (19%) participants had at least one PCR-confirmed positive result. Including re-infections, we observed 551 PCR-positive infections over 407,226 person-days of follow-up, corresponding to a crude (unadjusted) PCR positivity rate of 13.5 (95% confidence interval 12.4 to 14.7) per 10,000 days follow-up. Crude PCR positivity rates were higher for those with a waned booster vs. those who received a booster in October 2023 (15.3 (13.7 to 17.0) per 10,000 days follow-up vs. 11.5 (10.0 to 13.1)), and lower among those with a recent prior infection within 6 months vs. an infection 2+ years prior (6.7 (5.1 to 8.6) per 10,000 days follow-up vs. 14.2 (12.1 to 16.6)).

### Vaccine effectiveness

Relative to a waned booster, VE against infection for an October 2023 booster was estimated as 27.2% (95% confidence interval 10.6 to 40.7%) over the 6-month study period. VE was 40.5% (18.6 to 56.5%) at 0-2 months post-vaccination, 19.9% (-3.9 to 38.2%) at 2-4 months, and 13.8% (-41.5 to 47.5%) at 4-6 months (Table 2, Figure 2 panels A-B).

**Figure 2.**
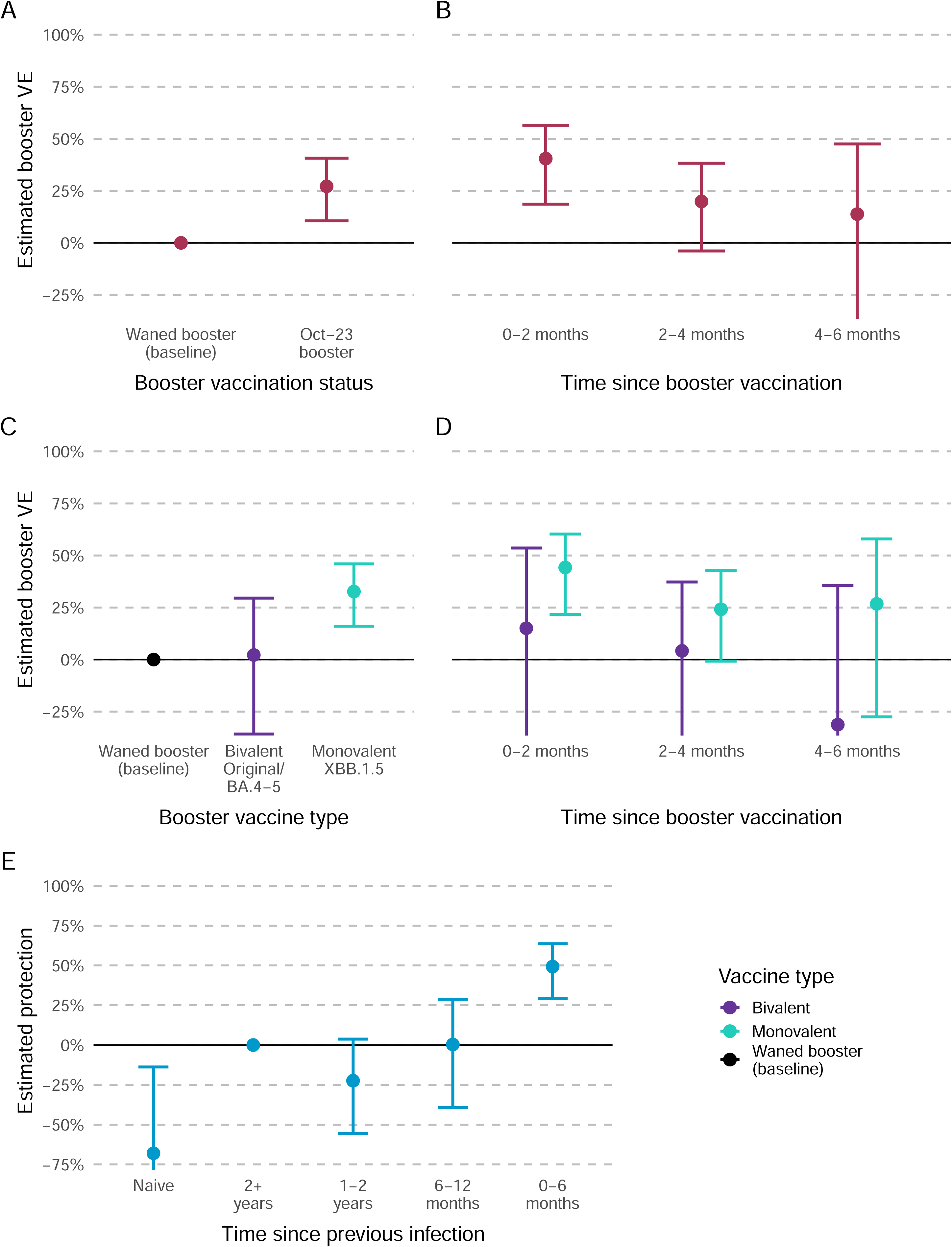

VE was 34.3% (-30.8 to 67.0%) among those confirmed as naïve, 37.1% (8.7 to 56.6%) for those with a previous infection more than 2 years ago, and 33.1% (9.4 to 50.6%) for those with an infection in the past 1-2 years (Table 3, Figure 3 panel A). Corresponding VE estimates by time since previous infection and booster vaccine type are shown in Figure 3 panel B.

**Figure 3.**
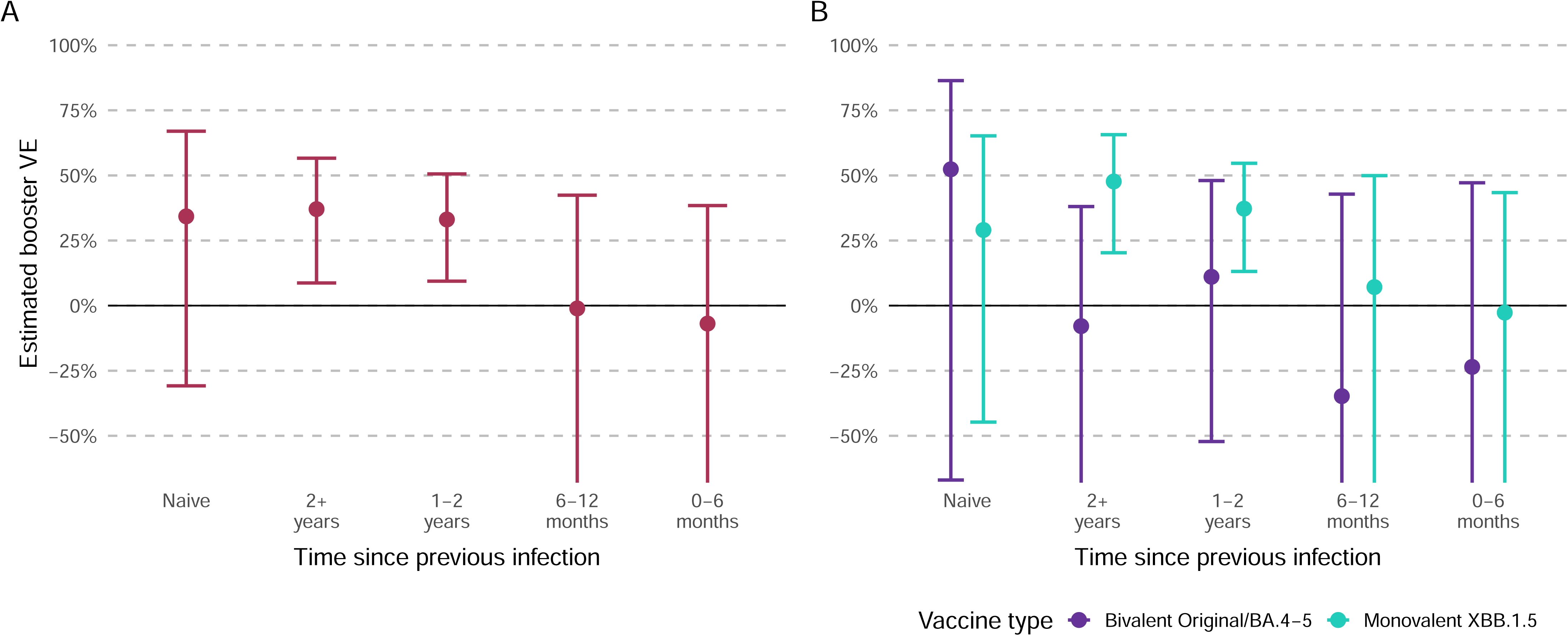

VE differed according to booster vaccine type; VE for the bivalent Original/BA.4-5 booster was 2.2% (-35.7 to 29.5%) overall, 15.1% (-55.4 to 53.6%) at 0-2 months, 4.2% (-46.4 to 37.3%) at 2-4 months, and -31.2% (-167.4 to 35.6%) at 4-6 months; VE for the Monovalent XBB.1.5 booster was 32.7% (16.1 to 46.0%) overall, 44.2% (21.7 to 60.3%) at 0-2 months, 24.1% (-0.7 to 42.9%) at 2-4 months, and 26.7% (-27.5 to 57.9%) at 4-6 months) (Table 2, Figure 2 panels C-D).

### Protection from previous infection

Based on estimates adjusted for month of study, vaccine status, and demographic covariates, compared to individuals with a COVID-19 infection more than 2 years ago, an infection within the past 6 months was associated with a 49.3% (29.2 to 63.6%) increase in protection, whereas those with no prior infection had -68.0% (-147.9 to -13.8%) reduced protection. An infection within the past 6-12 months, or an infection 1 to 2 years ago was not associated with significantly more or less protection compared to an infection more than 2 years ago; 0.3% (-39.3 to 28.7%) and -22.4% (-55.6 to 3.7%), respectively (Table 2, Figure 2 panel E).

### Moderate vs. mild/asymptomatic infection

Among the 551 positive PCR results, 543 had symptom information reported. Of these, 263 (48%) had moderate COVID-19 symptoms (ILI, sick leave, or ARI for 5+ days), whilst 280 (52%) had mild COVID-19 symptoms or were asymptomatic.

The proportion reporting moderate symptoms was lower among recipients of a booster dose (41%, 88/215), compared to those with no booster (52%, 175/336). Among those with an infection in the past 6 months, 33% (21/63) reported moderate symptoms compared to 50% (112/225) for those with an infection 1-2 years ago and 62% (28/45) for those with no previous infection (Table 2, Figure 4).

**Figure 4.**
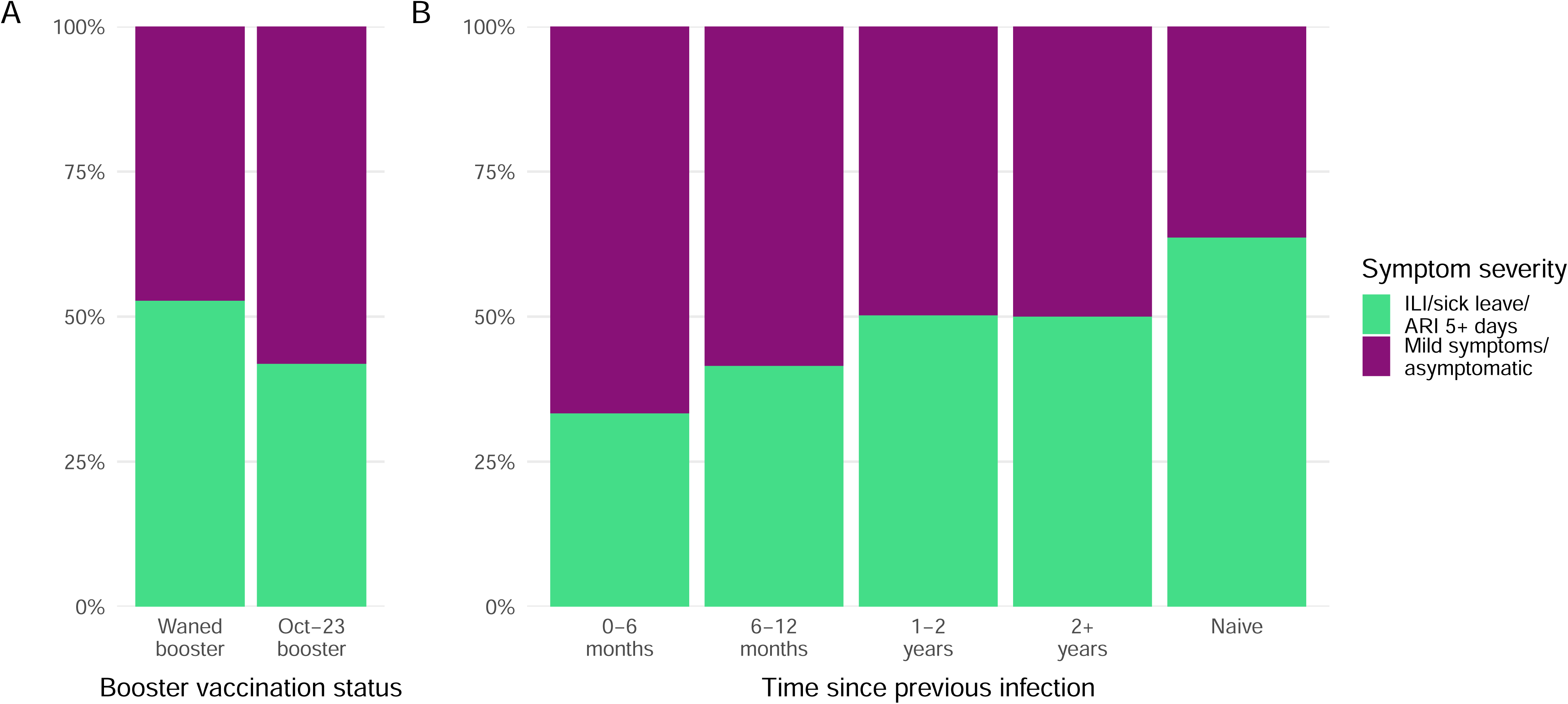

Relative to a waned booster, VE for an October 2023 booster was 39.7% (95% confidence interval 19.9 to 54.6%) against infection with moderate symptoms and 14.0% (-12.1 to 34.0%) against mild/asymptomatic infection (Table 2, Figure 5 panels A-B). A monovalent booster provided greater protection against infection with moderate symptoms compared to a bivalent booster (47.4% (27.9 to 61.6%) vs. 2.2% (-35.7 to 29.5%)). Protection declined over time but was more durable against moderate infection than against mild/asymptomatic infection: monovalent booster VE at 2-4 months was 36.8% (6.3 to 57.4%) against moderate infection compared to 12.0% (-26.4 to 38.8%) against mild infection (Table 2, Figure 5 panels C-D).

**Figure 5.**
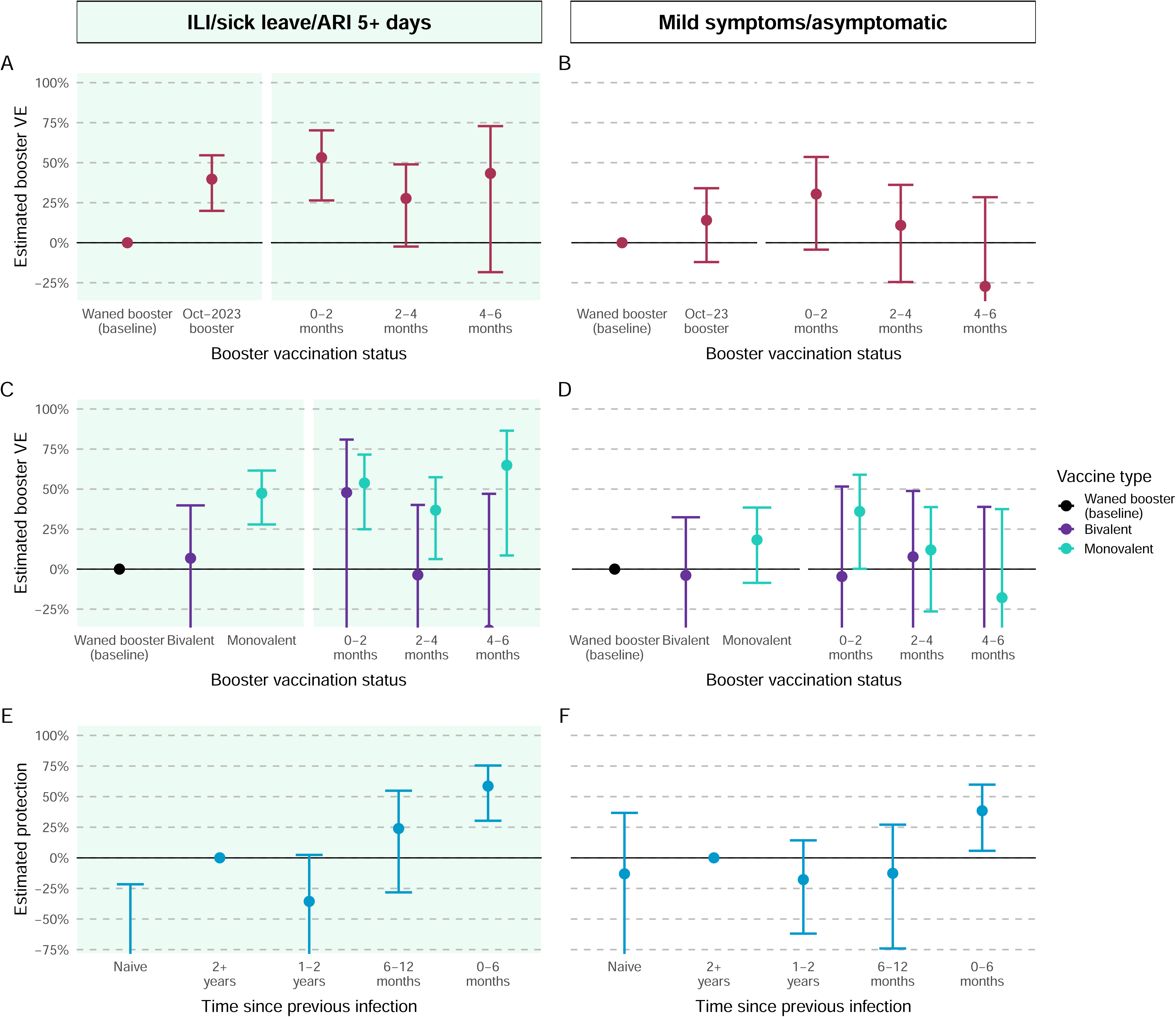

Relative to an infection 2+ years previously, an infection in the past 6 months was associated with 58.6% (30.3 to 75.4%) increased protection against moderate infection, and 38.5% (5.8 to 59.8%) increased protection against mild/asymptomatic infection (Table 2, Figure 5 panels E-F).

### Duration of PCR positivity

For a typical study participant (female, aged 45-54) receipt of a booster in October 2023 was associated with a reduced mean duration of PCR positivity; 7.7 days (5.3 to 11.2 days) vs. 11.1 days (95% confidence interval 7.9 to 15.5 days) among those with a waned booster (Table 4, Figure 6 panel A). Infections reported with moderate symptoms had a shorter duration of PCR positivity compared to mild infections; 9.9 days (6.8 to 14.5 days) vs. 12.3 days (8.4 to 18.0 days) for those with a waned booster, and 6.1 days (3.8 to 9.8 days) vs. 9.7 days (6.4 to 14.7 days) for those who received an October 2023 booster (Table 4, Figure 6 panel B). The estimated duration of PCR positivity was shorter for an individual with a recent prior infection; 5.5 days (3.6 to 8.4 days) for those with a prior infection within 6 months but 10.9 days (6.9 to 17.2 days) for prior infection within 6-12 months and 16.2 days (9.4 to 27.9 days) for those with a confirmed naïve infection status (Table 4, Figure 6 panel C). The differences in mean duration of PCR positivity by vaccination and prior infection were estimated for all participant groups (examples of other groups shown in Figures S9-S10).

**Figure 6.**
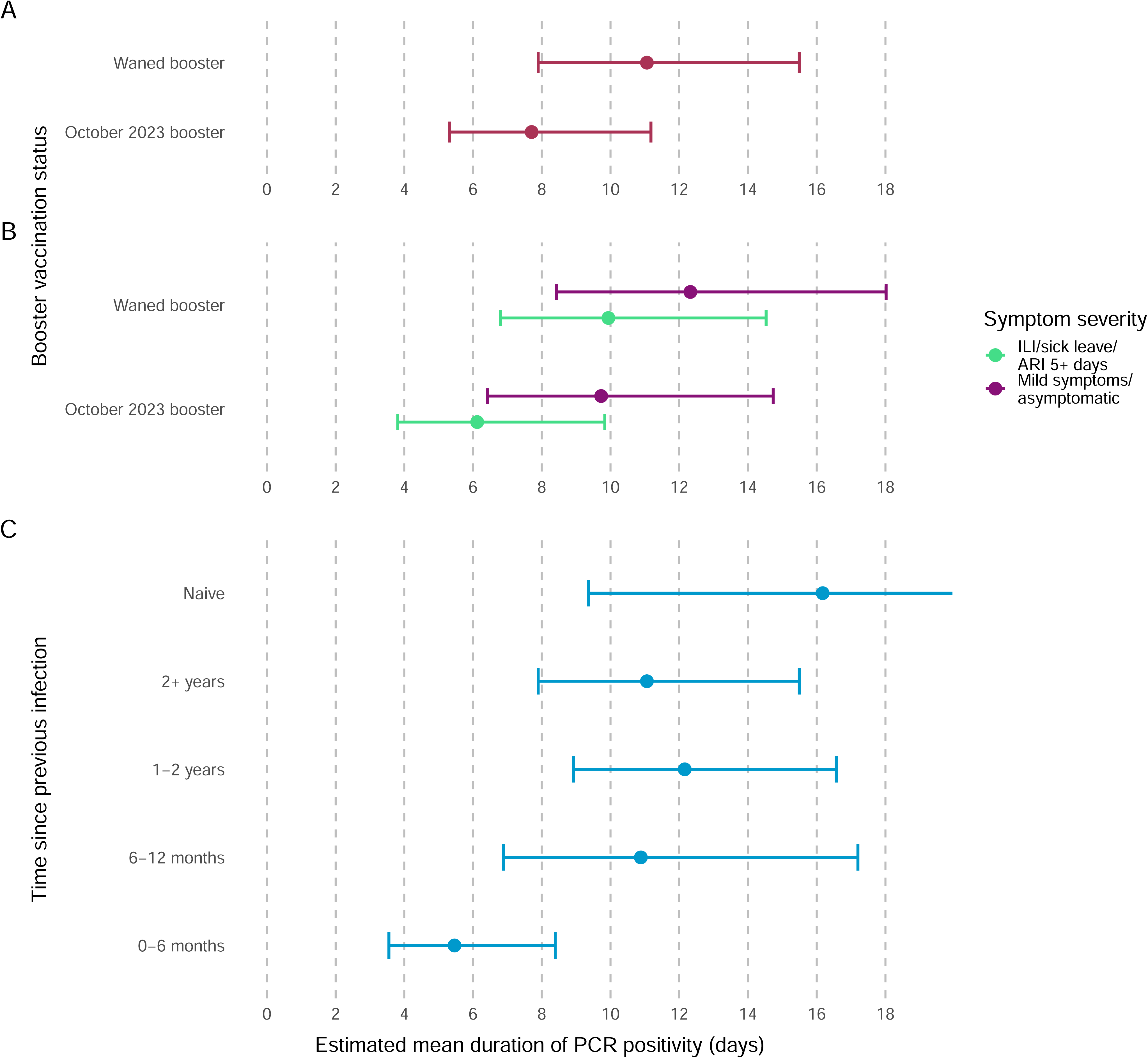

## Discussion

We have estimated COVID-19 booster vaccine effectiveness and protection following prior SARS-CoV-2 infection in a highly vaccinated and infection-experienced cohort of healthcare workers during autumn/winter 2023/24, a period of Omicron XBB.1.5 and JN.1 sub-variant circulation. We found that COVID-19 boosters provided moderate short-term protection against infection, compared to a waned booster (>6 months prior), with greater protection against moderate symptoms. Comparing recipients of the bivalent original/BA.4-5 and monovalent XBB.1.5 mRNA vaccine formulations, we estimated greater and longer-lasting protection from the monovalent booster, particularly against infection with moderate symptoms. As in previous analyses of this cohort [9,10], a recent prior infection was found to provide more sustained protection against infection compared with booster vaccination, with waning of this protection after 6 months. Controlling for vaccination, participants with a recent prior infection were less likely to experience moderate symptoms compared to those whose prior infection was more historic.

Our results are the most up-to-date evidence on the real-world effectiveness of two different COVID-19 booster formulations and have been interpreted in the context of high population immunity from previous infection and the ongoing evolution of SARS-CoV-2 Omicron sub-variants. An earlier study of symptomatic infection in the Netherlands estimated monovalent XBB.1.5 booster VE among working-age adults as 41% (23 to 55%), in line with our estimates for moderate infection [19]. Meanwhile, the improved real-world VE of the monovalent XBB.1.5 vaccine compared to the bivalent original/BA.4-5 vaccine supports findings from a comparable population-based cohort study in Singapore which estimated VE against symptomatic infection of 41% (34 to 48%) for the monovalent booster, compared to 8% (5 to 12%) for the bivalent booster, during a period of JN.1 transmission [18]. For previously uninfected individuals, markedly higher increases in mean neutralising antibody titres were seen following administration of a monovalent compared to a bivalent booster, with less severe immunological imprinting [20].

The multi-state models we fitted provide an estimate of the duration of PCR positivity. The estimates were in-line with other studies of Omicron BA.4/5 and JN.1 PCR positivity and viral shedding (around 8-10 days) [21], with differences between subgroups. Participants with asymptomatic/mild infection were estimated to have longer durations of PCR positivity than those with moderate illness. A study in China similarly estimated longer mean viral shedding periods for those with asymptomatic Omicron BA.5.2 COVID-19 infection compared to symptomatic [22]. Booster recipients had a shorter point estimate for duration of PCR positivity than those with a waned booster. We did not collect information on infectiousness, but a recent evidence review reported that COVID-19 cases were most infectious for 5 days after symptom onset, with most transmission events occurring during this time [21].

### Strengths and limitations

Two key strengths of our study are the ability to discern moderate and mild/asymptomatic infection based on high-quality symptoms reporting, and participants’ adherence to a regular testing regime. Half of detected infections were reported with moderate symptoms, including influenza-like illness, persistent acute respiratory symptoms, or requiring participants to take sick leave. Both a recent prior infection and receipt of a monovalent booster provided protection against moderate symptoms, as well as reducing the duration of PCR positivity. The continued follow-up of study participants for 3-years prior to the current analysis period (with regular PCR and antibody testing and access to complete COVID-19 vaccination history) adds confidence to these estimates, minimising the impact of reporting and recall bias [23].

The SIREN cohort is a cohort of working-age healthcare staff, with participants being predominantly female, of white ethnicity, healthy, and middle-aged. Whilst we control for these important factors in our analyses, differences in vaccination history, and the low proportion of males, older participants, and those with high multimorbidity limits generalisability of these findings to a wider UK population.

Another limitation is that booster vaccination was not randomly assigned, and despite limited differences in vaccine uptake by measured demographics, we could not control for several other prognostic factors which may be associated with vaccination, for example, an individual’s perceived exposure risk, which may alter their decision to receive a booster.

We did not investigate severe disease requiring hospitalisation, which was rare in this cohort (<5 participants self-reported hospitalisation for COVID-19 symptoms). For those aged 65 and older, a test-negative case-control study in England during September 2023 to January 2024 (a period of XBB.1.5 and JN.1 sub-variant circulation) estimated good protection of both the monovalent and bivalent boosters against hospitalisation, with VE point estimates being generally higher for the monovalent compared to the bivalent booster [24].

Given the very small number of unvaccinated individuals in SIREN and recognising they may have different risk profiles to vaccinated individuals, we were unable to consider them as a reference group to estimate absolute VE.

Finally, whilst the SIREN cohort is one of the most frequently tested COVID-19 cohorts still active, due to gaps in PCR testing and serum collection, including a pause in study testing between April and September 2023, we may have missed prior infections in the cohort. This resulted in the exclusion of 703 individuals whose infection status could not be ascertained due to gaps in their testing records.

### Implications of the study and future research

In this highly vaccinated, infection-experienced, working-age cohort there was a moderate but short-lived increase in protection against SARS-CoV-2 infection associated with receipt of a monovalent XBB.1.5 booster during the Omicron XBB.1.5 and JN.1 sub-variant circulating period. As new variants continue to emerge, ongoing monitoring of COVID-19 vaccine effectiveness is required to ensure the appropriateness and cost-effectiveness of vaccine programmes. Updated formulations which target the circulating variant are likely to be most effective in seasonal vaccination campaigns, however the lower coverage of COVID-19 boosters among healthcare workers, in comparison to seasonal influenza vaccination, limits their effectiveness at a population level.

## Supporting information

Figure headings

SIREN Study Group

## Supplementary details

### Contributors and guarantor information

VJH and SH conceived and supervised the study. PDK, SF, and VJH designed the analysis plan. PDK did the literature search and drafted the manuscript. PDK and SF cleaned and finalised the dataset for analysis and verified the data in the study. PDK planned and undertook the statistical analysis, with support from AC, AP, DDA, CHJ, and SS. VJH, SF, KM, DS, JS, AH, AD, JT, SR, JK, AA, and JI were responsible for SIREN 2.0 study delivery, participant recruitment and retention, data collection, data management, and quality improvement. ADO, DB, NH, and MDC were responsible for serum sampling and serological testing on samples collected 2023/24, which were used to inform categorisation of previous infection status. VJH, SF, KM, DS, JS, AH, AD, JT, SR, JK, DB, ADO, NH, MDC, CHJ, SS, CSB, AA, JI, AC, DDA, AMP, and SH revised the manuscript for important intellectual content and approved the manuscript for publication. PDK, SF, VJH, and SH had full access to all the data in the study, all authors accept responsibility to submit for publication. SH is the Chief Investigator of SIREN.

The corresponding author attests that all listed authors meet authorship criteria and that no others meeting the criteria have been omitted.

### Copyright/licence for publication

For the purpose of open access, the author has applied a Crown Copyright and Open Government Licence to any Author Accepted Manuscript version arising.

## Data sharing statement

Anonymised data will be made available for secondary analysis to trusted researchers upon reasonable request.

## Acknowledgements

We thank all the participants for their ongoing contributions and commitment to this study; the research teams at all 89 SIREN sites actively following up participants during this analysis period for their support and for making the study possible; and colleagues at the UK Health Security Agency Porton Down for organising and performing all the centralised antibody testing. We thank Diane Corrigan at the Public Health Agency Northern Ireland, Kevin Wilson at Public Health Scotland and Elen de Lacy in Public Health Wales and Chris Norman at Health and Care Research Wales for supporting SIREN 2.0 delivery and providing data from their national immunisation registries.

## Ethics approval

The study protocol was approved by the Berkshire Research Ethics Committee on May 22, 2020 (IRAS ID 284460, REC reference 20/SC/0230). The ethics amendment for testing over 2023/24 “SIREN 2.0” was approved on 14/08/2023 (Amendment 31). This study collected consent from all participants. ISRCTN Registry number: ISRCTN11041050.

## Dissemination to participants and related patient and public communities

We use diverse channels to disseminate our findings to participants, including via regular digital newsletters, live webinars, videos, blogs and through our study webpage on the UKHSA website (https://www.gov.uk/guidance/siren-study).

UKHSA and the MRC Biostatistics Unit have public facing websites and X accounts @UKHSA and @MRC_BSU. UKHSA and the MRC Biostatistics Unit engage with print and internet press, television, radio, news, and documentary programme makers.

## Transparency statement

The lead author affirms that this manuscript is an honest, accurate, and transparent account of the study being reported; that no important aspects of the study have been omitted; and that any discrepancies from the study as planned (and, if relevant, registered) have been explained.

## Data availability

The anonymised SIREN dataset will be made available for secondary analysis to approved researchers upon reasonable request, subject to appropriate information governance requirements.

## Code availability

A list of all R packages used, and R code written to process the data, implement the statistical analysis, and produce the figures and tables is available online at: https://github.com/SIREN-study/SARS-CoV-2-third-booster.

