## Supplementary material for "Protection of vaccine boosters and prior infection against mild/asymptomatic and moderate COVID-19 infection in the UK SIREN healthcare worker cohort: October 2023 to March 2024": Tables

Table 1: Demographic characteristics of SIREN study participants, by receipt of COVID-19 booster in October 2023.

|  | | **Receipt of COVID-19 booster** | |
| --- | --- | --- | --- |
| **Characteristic** | **Overall**  N = 2,867 | **Waned booster** N = 1,445 | **October 2023 booster**  N = 1,422 |
| Gender |  |  |  |
| Female | 2,311 (80.6%) | 1,214 (84.0%) | 1,097 (77.1%) |
| Male | 556 (19.4%) | 231 (16.0%) | 325 (22.9%) |
| Age group |  |  |  |
| Under 35 | 195 (6.80%) | 117 (8.10%) | 78 (5.49%) |
| 35 to 44 | 638 (22.3%) | 342 (23.7%) | 296 (20.8%) |
| 45 to 54 | 1,149 (40.1%) | 579 (40.1%) | 570 (40.1%) |
| 55 to 64 | 822 (28.7%) | 389 (26.9%) | 433 (30.5%) |
| Over 65 | 63 (2.20%) | 18 (1.25%) | 45 (3.16%) |
| Ethnicity |  |  |  |
| White | 2,450 (85.5%) | 1,193 (82.6%) | 1,257 (88.4%) |
| Asian | 223 (7.78%) | 128 (8.86%) | 95 (6.68%) |
| Black | 90 (3.14%) | 59 (4.08%) | 31 (2.18%) |
| Mixed | 51 (1.78%) | 33 (2.28%) | 18 (1.27%) |
| Other | 53 (1.85%) | 32 (2.21%) | 21 (1.48%) |
| Medical condition |  |  |  |
| No medical condition | 2,117 (73.8%) | 1,097 (75.9%) | 1,020 (71.7%) |
| Immunosuppression | 70 (2.44%) | 30 (2.08%) | 40 (2.81%) |
| Chronic Respiratory conditions | 342 (11.9%) | 161 (11.1%) | 181 (12.7%) |
| Chronic Non-Respiratory conditions | 338 (11.8%) | 157 (10.9%) | 181 (12.7%) |
| Staff type |  |  |  |
| Administrative/Executive (office based) | 466 (16.3%) | 224 (15.5%) | 242 (17.0%) |
| Doctor | 366 (12.8%) | 143 (9.90%) | 223 (15.7%) |
| Estates/Porters/Security | 53 (1.85%) | 29 (2.01%) | 24 (1.69%) |
| Healthcare Assistant | 155 (5.41%) | 96 (6.64%) | 59 (4.15%) |
| Healthcare Scientist | 183 (6.38%) | 97 (6.71%) | 86 (6.05%) |
| Midwife | 91 (3.17%) | 59 (4.08%) | 32 (2.25%) |
| Nursing | 952 (33.2%) | 508 (35.2%) | 444 (31.2%) |
| Other | 334 (11.6%) | 167 (11.6%) | 167 (11.7%) |
| Pharmacist | 64 (2.23%) | 25 (1.73%) | 39 (2.74%) |
| Physiotherapist/Occupational Therapist/SALT | 122 (4.26%) | 62 (4.29%) | 60 (4.22%) |
| Student (Medical/Nursing/Midwifery/Other) | 81 (2.83%) | 35 (2.42%) | 46 (3.23%) |
| Occupation setting |  |  |  |
| Ambulance/Emergency Department | 57 (1.99%) | 29 (2.01%) | 28 (1.97%) |
| Inpatient Wards | 350 (12.2%) | 207 (14.3%) | 143 (10.1%) |
| Intensive Care | 128 (4.46%) | 69 (4.78%) | 59 (4.15%) |
| Maternity/Labour Ward | 45 (1.57%) | 29 (2.01%) | 16 (1.13%) |
| Office | 629 (21.9%) | 282 (19.5%) | 347 (24.4%) |
| Outpatient | 585 (20.4%) | 302 (20.9%) | 283 (19.9%) |
| Patient facing (non-clinical) | 136 (4.74%) | 62 (4.29%) | 74 (5.20%) |
| Theatres | 75 (2.62%) | 42 (2.91%) | 33 (2.32%) |
| Other | 862 (30.1%) | 423 (29.3%) | 439 (30.9%) |
| Patient contact |  |  |  |
| Yes | 2,411 (84.1%) | 1,233 (85.3%) | 1,178 (82.8%) |
| No | 456 (15.9%) | 212 (14.7%) | 244 (17.2%) |
| Index of multiple deprivation |  |  |  |
| Most deprived (1) | 255 (9.00%) | 152 (10.6%) | 103 (7.35%) |
| Deprivation 2 | 470 (16.6%) | 265 (18.5%) | 205 (14.6%) |
| Deprivation 3 | 606 (21.4%) | 314 (21.9%) | 292 (20.8%) |
| Deprivation 4 | 721 (25.5%) | 348 (24.3%) | 373 (26.6%) |
| Least deprived (5) | 781 (27.6%) | 353 (24.7%) | 428 (30.5%) |
| Unknown | 34 | 13 | 21 |
| Region |  |  |  |
| East Midlands | 283 (9.87%) | 128 (8.86%) | 155 (10.9%) |
| East of England | 273 (9.52%) | 131 (9.07%) | 142 (9.99%) |
| London | 374 (13.0%) | 205 (14.2%) | 169 (11.9%) |
| North East | 51 (1.78%) | 13 (0.90%) | 38 (2.67%) |
| North West | 333 (11.6%) | 179 (12.4%) | 154 (10.8%) |
| Scotland | 417 (14.5%) | 247 (17.1%) | 170 (12.0%) |
| South East | 320 (11.2%) | 151 (10.4%) | 169 (11.9%) |
| South West | 309 (10.8%) | 126 (8.72%) | 183 (12.9%) |
| West Midlands | 216 (7.53%) | 148 (10.2%) | 68 (4.78%) |
| Yorkshire and the Humber | 291 (10.1%) | 117 (8.10%) | 174 (12.2%) |
| Household structure |  |  |  |
| Lives with others (including children) | 1,062 (37.0%) | 573 (39.7%) | 489 (34.4%) |
| Lives with others (no children) | 1,436 (50.1%) | 708 (49.0%) | 728 (51.2%) |
| Lives alone | 369 (12.9%) | 164 (11.3%) | 205 (14.4%) |
| Time since previous infection (at study entry) |  |  |  |
| Naïve | 166 (5.79%) | 58 (4.01%) | 108 (7.59%) |
| 2+ years | 786 (27.4%) | 416 (28.8%) | 370 (26.0%) |
| 1-2 years | 1,033 (36.0%) | 538 (37.2%) | 495 (34.8%) |
| 6-12 months | 424 (14.8%) | 231 (16.0%) | 193 (13.6%) |
| 0-6 months | 458 (16.0%) | 202 (14.0%) | 256 (18.0%) |
| Vaccine type |  |  |  |
| Waned booster | 1,445 (50.4%) | 1,445 (100.0%) | 0 (0%) |
| Bivalent Original/BA.4-5 | 280 (9.77%) | 0 (0%) | 280 (19.7%) |
| Monovalent XBB.1.5 | 1,142 (39.8%) | 0 (0%) | 1,142 (80.3%) |

Table 2: Crude PCR positivity rates per 10,000 person-days and estimated vaccine effectiveness and protection from prior infection by booster vaccine status, time since booster, booster vaccine type, time since previous infection, and COVID-19 symptom severity.

|  | **Number of participants** | **Positive PCR tests** | **Exposure (person-days at risk)** | **Crude PCR positivity rate per 10,000 person-days (95% CI)** | **Protection relative to baseline (95% CI)** | **ILI/sick leave/ARI 5+ days** | | **Mild symptoms/ asymptomatic** | |
| --- | --- | --- | --- | --- | --- | --- | --- | --- | --- |
|  |  |  |  |  |  | **Positive PCR tests** | **Protection (VE) relative to baseline (95% CI)** | **Positive PCR tests** | **Protection (VE) relative to baseline (95% CI)** |
| Whole population | 2,867 | 551 | 407,226 | 13.5 (12.4, 14.7) | N/A | 263 | N/A | 280 | N/A |
| **Booster vaccine status** | | |  |  |  |  |  |  |  |
| Waned booster | 2,791 | 336 | 219,849 | 15.3 (13.7, 17.0) | Baseline | 175 | Baseline | 156 | Baseline |
| October 2023 booster* | 1,421 | 215 | 187,377 | 11.5 (10.0, 13.1) | 27.2% (10.6, 40.7) | 88 | 39.7% (19.9, 54.6) | 124 | 14.0% (-12.1, 34.0) |
| **Time since booster** | |  |  |  |  |  |  |  |  |
| 0-2 months | 1,369 | 47 | 56,279 | 8.4 (6.1, 11.1) | 40.5% (18.6, 56.5) | 18 | 53.2% (26.4, 70.2) | 29 | 30.4% (-4.3, 53.6) |
| 2-4 months | 1,383 | 119 | 75,340 | 15.8 (13.1, 18.9) | 19.9% (-3.9, 38.2) | 54 | 27.7% (-2.4, 48.9) | 63 | 10.8% (-24.5, 36.2) |
| 4-6 months | 1,337 | 49 | 55,758 | 8.8 (6.5, 11.6) | 13.8% (-41.5, 47.5) | 16 | 43.3% (-18.4, 72.9) | 32 | -27.3% (-126.3, 28.4) |
| **Booster vaccine type** | |  |  |  |  |  |  |  |  |
| Bivalent Original/BA.4-5 | 280 | 55 | 37,587 | 14.6 (11.0, 19.0) | 2.2% (-35.7, 29.5) | 25 | 6.8% (-44.3, 39.8) | 29 | -3.9% (-59.7, 32.4) |
| Monovalent XBB.1.5 | 1,141 | 160 | 149,790 | 10.7 (9.1, 12.5) | 32.7% (16.1, 46.0) | 63 | 47.4% (27.9, 61.6) | 95 | 18.3% (-8.6, 38.5) |
| **Booster vaccine type and time since booster** | | | | | |  |  |  |  |
| Bivalent 0-2 months | 269 | 10 | 11,037 | 9.1 (4.3, 16.7) | 15.1% (-55.4, 53.6) | 2 | 47.8% (-42.6, 80.9) | 8 | -4.6% (-126.2, 51.6) |
| Bivalent 2-4 months | 271 | 31 | 14,690 | 21.1 (14.3, 30.0) | 4.2% (-46.4, 37.3) | 16 | -3.7% (-79.4, 40.1) | 14 | 7.7% (-66.4, 48.8) |
| Bivalent 4-6 months | 259 | 14 | 11,860 | 11.8 (6.5, 19.8) | -31.2% (-167.4, 35.6) | 7 | -38.1% (-260.1, 47.0) | 7 | -53.9% (-287.6, 38.9) |
| Monovalent 0-2 months | 1,100 | 37 | 45,242 | 8.2 (5.8, 11.3) | 44.2% (21.7, 60.3) | 16 | 53.8% (24.9, 71.6) | 21 | 36.0% (0.2, 59.0) |
| Monovalent 2-4 months | 1,112 | 88 | 60,650 | 14.5 (11.6, 17.9) | 24.1% (-0.7, 42.9) | 38 | 36.8% (6.3, 57.4) | 49 | 12.0% (-26.4, 38.8) |
| Monovalent 4-6 months | 1,078 | 35 | 43,898 | 8.0 (5.6, 11.1) | 26.7% (-27.5, 57.9) | 9 | 64.8% (8.5, 86.5) | 25 | -17.8% (-122.1, 37.5) |
| **Time since previous infection** | | |  |  |  |  |  |  |  |
| Naïve | 166 | 45 | 20,764 | 21.7 (15.8, 29.0) | -68.0% (-147.9, -13.8) | 28 | -101.2% (-233.0, -21.6) | 16 | -13.0% (-102.1, 36.8) |
| 2+ years | 1,058 | 163 | 114,534 | 14.2 (12.1, 16.6) | Baseline | 80 | Baseline | 81 | Baseline |
| 1-2 years | 1,303 | 225 | 134,964 | 16.7 (14.6, 19.0) | -22.4% (-55.6, 3.7) | 112 | -35.6% (-88.3, 2.4) | 110 | -17.8% (-61.9, 14.3) |
| 6-12 months | 545 | 55 | 42,696 | 12.9 (9.7, 16.8) | 0.3% (-39.3, 28.7) | 22 | 23.9% (-28.2, 54.9) | 32 | -12.6% (-74.1, 27.1) |
| 0-6 months | 912 | 63 | 94,268 | 6.7 (5.1, 8.6) | 49.3% (29.2, 63.6) | 21 | 58.6% (30.3, 75.4) | 41 | 38.5% (5.8, 59.8) |

*n=1 boosted participant contributed no follow-up time post-booster receipt.

Table 3: Estimated booster vaccine effectiveness (VE) relative to a waned booster, by time since previous infection and booster vaccine type.

| Time since previous infection | October 2023 booster (overall)  (95% CI) | Bivalent Original/BA.4-5  (95% CI) | Monovalent XBB.1.5  (95% CI) |
| --- | --- | --- | --- |
| Naive | 34.3% (-30.8, 67.0) | 52.4% (-67.0, 86.4) | 29.0% (-44.7, 65.2) |
| 2+ years | 37.1% (8.7, 56.6) | -7.9% (-87.8, 38.0) | 47.7% (20.3, 65.6) |
| 1-2 years | 33.1% (9.4, 50.6) | 11.1% (-52.2, 48.0) | 37.2% (13.1, 54.6) |
| 6-12 months | -1.1% (-77.4, 42.4) | -34.8% (-217.5, 42.8) | 7.1% (-72.3, 49.9) |
| 0-6 months | -6.9% (-85.6, 38.4) | -23.5% (-189.0, 47.2) | -2.7% (-86.3, 43.4) |

Table 4: Estimated mean duration of PCR positivity (days) for a typical study participant (female, aged 45-54), by vaccination status, time since previous infection and symptom severity.

|  | Estimated mean duration of PCR positivity (95% CI) |
| --- | --- |
| Waned booster dose | 11.1 days (7.9, 15.5) |
| October 2023 booster | 7.7 days (5.3, 11.2) |
| Waned booster dose |  |
| ILI/sick leave/symptoms 5+ days | 9.9 days (6.8, 14.5) |
| Mild symptoms/asymptomatic | 12.3 days (8.4, 18.0) |
| October 2023 booster |  |
| ILI/sick leave/symptoms 5+ days | 6.1 days (3.8, 9.8) |
| Mild symptoms/asymptomatic | 9.7 days (6.4, 14.7) |
| Naïve | 16.2 days (9.4, 27.9) |
| 2+ years | 11.1 days (7.9, 15.5) |
| 1-2 years | 12.2 days (8.9, 16.6) |
| 6-12 months | 10.9 days (6.9, 17.2) |
| 0-6 months | 5.5 days (3.6, 8.4) |
