## Supplementary material for "Protection of vaccine boosters and prior infection against mild/asymptomatic and moderate COVID-19 infection in the UK SIREN healthcare worker cohort: October 2023 to March 2024": Figure headings

Figure 1: Consort diagram showing participation in study and uptake of booster vaccine.

Figure 2: Estimated booster vaccine effectiveness (VE), relative to waned booster, by booster vaccine status (panel A, model M1), time since booster vaccination (panel B, model M2), and booster vaccine type (panels C and D, models M3 and M4), and estimated protection from previous infection, relative to a baseline of 2+ years (panel E, model M2). Error bars show the 95% confidence intervals.

Figure 3: Estimated booster vaccine effectiveness (VE) relative to a waned booster, by time since previous infection (panel A, model M1), and time since previous infection and booster vaccine type (panel B, model M3). Error bars show the 95% confidence intervals.

Figure 4: SARS-CoV-2 infections by COVID-19 symptom severity and booster vaccine status (panel A) and time since previous infection (panel B).

Figure 5: Estimated booster vaccine effectiveness (VE) by symptom severity, relative to waned booster dose, by booster vaccination status and time since booster vaccination (panels A and B, models M5 and M6), and vaccine type (panels C and D, models M7 and M8) and estimated protection from previous infection, relative to a baseline of 2+ years (panels E and F, model M5). Panels with shaded background indicate VE against moderate symptoms. Error bars show the 95% confidence intervals.

Figure 6: Estimated mean duration of PCR positivity (days) for a typical study participant (female, aged 45-54), by vaccination status (panel A, model M1), vaccination status and symptom severity (panel B, model M4), and time since previous infection (panel C, model M1). Error bars show the 95% confidence intervals for the mean duration of PCR positivity.
