## Supplementary material for "Protection of vaccine boosters and prior infection against mild/asymptomatic and moderate COVID-19 infection in the UK SIREN healthcare worker cohort: October 2023 to March 2024": SIREN Study Group

| **Organisation Name** | **First Name/Initial** | **Surname** |
| --- | --- | --- |
| **Participating NHS Sites for SIREN 2.0** |  |  |
| ANEURIN BEVAN UNIVERSITY LHB | John | Northfield |
| ANEURIN BEVAN UNIVERSITY LHB | Sean | Cutler |
| ASHFORD AND ST PETER'S HOSPITALS NHS FOUNDATION TRUST | Stephen | Winchester |
| ASHFORD AND ST PETER'S HOSPITALS NHS FOUNDATION TRUST | Samuel | Rowley |
| BELFAST HEALTH & SOCIAL CARE TRUST | Clodagh | Loughrey |
| BLACK COUNTRY HEALTHCARE NHS FOUNDATION TRUST | Alison | Grant |
| BLACK COUNTRY HEALTHCARE NHS FOUNDATION TRUST | Rebecca | Temple-Purcell |
| BUCKINGHAMSHIRE HEALTHCARE NHS TRUST | Nick | Wong |
| BUCKINGHAMSHIRE HEALTHCARE NHS TRUST | Ruth | Penn |
| CENTRAL AND NORTH WEST LONDON NHS FOUNDATION TRUST | Abigail | Severn |
| CENTRAL AND NORTH WEST LONDON NHS FOUNDATION TRUST | Alejandro | Arenas-Pinto |
| CWM TAF MORGANNWG UNIVERSITY LHB | John | Geen |
| CWM TAF MORGANNWG UNIVERSITY LHB | Carla | Pothecary |
| DERBYSHIRE HEALTHCARE NHS FOUNDATION TRUST | Joely | Morgan |
| DERBYSHIRE HEALTHCARE NHS FOUNDATION TRUST | Gemma | Harrison |
| GOLDEN JUBILEE NATIONAL HOSPITAL | Catherine | Sinclair |
| GOLDEN JUBILEE NATIONAL HOSPITAL | Val | Irvine |
| HOUNSLOW AND RICHMOND COMMUNITY HEALTHCARE NHS TRUST | John | Omany |
| HOUNSLOW AND RICHMOND COMMUNITY HEALTHCARE NHS TRUST | Shekoo | Mackay |
| ISLE OF WIGHT NHS TRUST | Emily | Macnaughton |
| ISLE OF WIGHT NHS TRUST | Sarah | Knight |
| JAMES PAGET UNIVERSITY HOSPITALS NHS FOUNDATION TRUST | Davis | Nwaka |
| JAMES PAGET UNIVERSITY HOSPITALS NHS FOUNDATION TRUST | Christian | Hacon |
| LANCASHIRE & SOUTH CUMBRIA NHS FOUNDATION TRUST | Robert | Shorten |
| LANCASHIRE & SOUTH CUMBRIA NHS FOUNDATION TRUST | Kathryn | Hollinshead |
| LEEDS TEACHING HOSPITALS NHS TRUST | Jacqueline | Brandon |
| LEEDS TEACHING HOSPITALS NHS TRUST | Kyra | Holliday |
| LIVERPOOL UNIVERSITY HOSPITALS NHS FOUNDATION TRUST | Anu | Chawla |
| LIVERPOOL UNIVERSITY HOSPITALS NHS FOUNDATION TRUST | Fran | Westwell |
| MANCHESTER UNIVERSITY NHS FOUNDATION TRUST | Alexander | Horsley |
| MANCHESTER UNIVERSITY NHS FOUNDATION TRUST | Shazaad | Ahmad |
| MID CHESHIRE HOSPITALS NHS FOUNDATION TRUST | Elijah | Matovu |
| MID CHESHIRE HOSPITALS NHS FOUNDATION TRUST | Claire | Gabriel |
| MID CHESHIRE HOSPITALS NHS FOUNDATION TRUST | Sheron | Clarke |
| NHS FIFE | Devesh | Dhasmana |
| NHS FIFE | Susan | Fowler |
| NHS FORTH VALLEY | Euan | Cameron |
| NHS FORTH VALLEY | Anne | Todd |
| NHS GREATER GLASGOW AND CLYDE | Antonia | Ho |
| NHS GREATER GLASGOW AND CLYDE | Michael | Murphy |
| NHS HIGHLAND | Andrew | Gibson |
| NHS HIGHLAND | Alexandra | Cochrane |
| NHS LANARKSHIRE | Manish | Patel |
| NHS LANARKSHIRE | Berni | Welsh |
| NHS LOTHIAN | Kate | Templeton |
| NHS LOTHIAN | Sam | Donaldson |
| NHS WESTERN ISLES | Martin | Malcolm |
| NHS WESTERN ISLES | Beth | Smith |
| NORFOLK AND NORWICH UNIVERSITY HOSPITALS NHS FOUNDATION TRUST | Ngozi | Elumogo |
| NORFOLK AND NORWICH UNIVERSITY HOSPITALS NHS FOUNDATION TRUST | Louise | Coke |
| NOTTINGHAM UNIVERSITY HOSPITALS NHS TRUST | Sarah | Brand |
| NOTTINGHAM UNIVERSITY HOSPITALS NHS TRUST | Jack | Squires |
| ROYAL PAPWORTH HOSPITAL NHS FOUNDATION TRUST | Sumita | Pai |
| ROYAL PAPWORTH HOSPITAL NHS FOUNDATION TRUST | Allison | Doel |
| SALISBURY NHS FOUNDATION TRUST | Abby | Rand |
| SALISBURY NHS FOUNDATION TRUST | Catherine | Thompson |
| SHEFFIELD CHILDREN'S NHS FOUNDATION TRUST | Fiona | Shackley |
| SHEFFIELD CHILDREN'S NHS FOUNDATION TRUST | James | Pethick |
| SHREWSBURY AND TELFORD HOSPITAL NHS TRUST | Mandy | Carnahan |
| SHREWSBURY AND TELFORD HOSPITAL NHS TRUST | Mandy | Beekes |
| SOUTH EASTERN HEALTH & SOCIAL CARE | Yuri | Protaschik |
| SOUTH EASTERN HEALTH & SOCIAL CARE | Susan | Regan |
| SOUTHERN HEALTH & SOCIAL CARE TRUST | Angel | Boulos |
| SOUTHERN HEALTH & SOCIAL CARE TRUST | Fiona | Thompson |
| ST GEORGE'S UNIVERSITY HOSPITALS NHS FOUNDATION TRUST | Tim | Planche |
| ST GEORGE'S UNIVERSITY HOSPITALS NHS FOUNDATION TRUST | Angela | Houston |
| STOCKPORT NHS FOUNDATION TRUST | Sharman | Harris |
| STOCKPORT NHS FOUNDATION TRUST | Barzo | Faris |
| THE NEWCASTLE UPON TYNE HOSPITALS NHS FOUNDATION TRUST | Brendan | Payne |
| THE NEWCASTLE UPON TYNE HOSPITALS NHS FOUNDATION TRUST | Jayne | Harwood |
| THE ROBERT JONES AND AGNES HUNT ORTHOPAEDIC HOSPITAL NHS FOUNDATION TRUST | Ruth | Longfellow |
| TORBAY AND SOUTH DEVON NHS FOUNDATION TRUST | Kelly | Barrett |
| TORBAY AND SOUTH DEVON NHS FOUNDATION TRUST | Matthew | Halkes |
| UNIVERSITY HOSPITALS BRISTOL AND WESTON NHS FOUNDATION TRUST | Rajeka | Lazarus |
| UNIVERSITY HOSPITALS BRISTOL AND WESTON NHS FOUNDATION TRUST | Aaran | Sinclair |
| UNIVERSITY HOSPITALS OF DERBY AND BURTON NHS FOUNDATION TRUST | L | Berry |
| UNIVERSITY HOSPITALS OF DERBY AND BURTON NHS FOUNDATION TRUST | Frances | Game |
| UNIVERSITY HOSPITALS OF LEICESTER NHS TRUST | Christopher | Holmes |
| UNIVERSITY HOSPITALS OF LEICESTER NHS TRUST | Martin | Wiselka |
| UNIVERSITY HOSPITALS OF MORECAMBE BAY NHS FOUNDATION TRUST | Timothy | Gatheral |
| UNIVERSITY HOSPITALS OF MORECAMBE BAY NHS FOUNDATION TRUST | Lynda | Fothergill |
| WHITTINGTON HEALTH NHS TRUST | Chetan | Parmar |
| WHITTINGTON HEALTH NHS TRUST | Philippa | Kemsley |
| **SIREN Study team** |  |  |
| UK HEALTH SECURITY AGENCY | Andre | Charlett |
| UK HEALTH SECURITY AGENCY | Omoyeni | Adebiyi |
| UK HEALTH SECURITY AGENCY | Nick | Andrews |
| UK HEALTH SECURITY AGENCY | Ana | Atti |
| UK HEALTH SECURITY AGENCY | Colin | Brown |
| UK HEALTH SECURITY AGENCY | Angela | Dunne |
| UK HEALTH SECURITY AGENCY | Sarah | Foulkes |
| UK HEALTH SECURITY AGENCY | Victoria | Hall |
| UK HEALTH SECURITY AGENCY | Nipunadi | Hettiarachchi |
| UK HEALTH SECURITY AGENCY | Susan | Hopkins |
| UK HEALTH SECURITY AGENCY | Anna | Howells |
| UK HEALTH SECURITY AGENCY | Jasmin | Islam |
| UK HEALTH SECURITY AGENCY | Jameel | Khawam |
| UK HEALTH SECURITY AGENCY | Katie | Munro |
| UK HEALTH SECURITY AGENCY | Sophie | Russell |
| UK HEALTH SECURITY AGENCY | Chantal | Sinclair |
| UK HEALTH SECURITY AGENCY | Dominic | Sparkes |
| UK HEALTH SECURITY AGENCY | Jean | Timeyin |
| UK HEALTH SECURITY AGENCY | Ashley | Otter |
| UK HEALTH SECURITY AGENCY | Cathy | Rowe |
| PUBLIC HEALTH AGENCY NORTHERN IRELAND | Diane | Corrigan |
| PUBLIC HEALTH AGENCY NORTHERN IRELAND | Lisa | Cromey |
| PUBLIC HEALTH SCOTLAND | Laura | Dobbie |
| PUBLIC HEALTH SCOTLAND | Kevin | Wilson |
| PUBLIC HEALTH WALES | Ellen | De Lacy |
| HEALTH AND CARE RESEARCH WALES | Chris | Norman |
| PUBLIC HEALTH WALES | Guy | Stevens |
| MRC BIOSTATISTICS UNIT, UNIVERSITY OF CAMBRIDGE | Peter | Kirwan |
| MRC BIOSTATISTICS UNIT, UNIVERSITY OF CAMBRIDGE | Christopher | Jackson |
| MRC BIOSTATISTICS UNIT, UNIVERSITY OF CAMBRIDGE | Anne | Presanis |
| MRC BIOSTATISTICS UNIT, UNIVERSITY OF CAMBRIDGE | Daniela | De Angelis |
| **SIREN 2.0 Academic Collaborators** |  |  |
| PROTECTIVE IMMUNITY FROM T CELLS TO COVID-19 IN HEALTH WORKERS (PITCH) STUDY, UNIVERSITY OF OXFORD | Susanna | Dunachie |
| PITCH Study, UNIVERSITY OF OXFORD | Paul | Klenerman |
| PITCH Study, UNIVERSITY OF NEWCASTLE | Chris | Duncan |
| PITCH Study, UNIVERSITY OF NEWCASTLE | Rebecca | Payne |
| PITCH Study, UNIVERSITY OF LIVERPOOL | Lance | Turtle |
| PITCH Study, UNIVERSITY OF BIRMINGHAM | Alex | Richter |
| PITCH Study, UNIVERSITY OF SHEFFIELD | Thushan | De Silva |
| PITCH Study, UNIVERSITY OF OXFORD | Eleanor | Barnes |
| PITCH Study, UNIVERSITY OF NEWCASTLE | Daniel | Wootton |
| THE FRANCIS CRICK INSTITUTE | Rupert | Beale |
| THE FRANCIS CRICK INSTITUTE | Edward | Carr |
| THE FRANCIS CRICK INSTITUTE | Mary | Wu |
| THE FRANCIS CRICK INSTITUTE | Ruth | Harvey |
| THE FRANCIS CRICK INSTITUTE | Nicola | Lewis |
| THE WELLCOME SANGER INSTITUTE | Ewan | Harrison |
| THE WELLCOME SANGER INSTITUTE | Ya-Lin | Huang |
| THE WELLCOME SANGER INSTITUTE | Katie | Bellis |
| THE WELLCOME SANGER INSTITUTE | Marissa | Knoll |
| BRITISH SOCIETY FOR IMMUNOLOGY | Jennie | Evans |
| BRITISH SOCIETY FOR IMMUNOLOGY | Hana | Ayoob |
